# Household Transmission of Enterovirus D68 in Washington and Oregon, USA, 2022-2024

**DOI:** 10.64898/2026.02.16.26346322

**Authors:** Pavitra Roychoudhury, Erica Wetzler, Anna Elias-Warren, Alex Harteloo, Hyeong Geon Kim, Kevin Kong, Hong Xie, Jolene Gov, Margaret G. Mills, Collrane Frivold, Madison Hollcroft, Mark Drummond, Tara Hatchie, Erica Clark, Brenna Ehmen, Peter D. Han, Luis Gamboa, Sally Grindstaff, Jeremy Stone, Alexander L. Greninger, Lea M. Starita, Christina Lockwood, Janet A. Englund, Ana A. Weil, Sacha L. Reich, Richard A. Mularski, Mark A Schmidt, Jennifer L. Kuntz, Allison L. Naleway, Helen Y. Chu

## Abstract

Household transmission of EV-D68 was identified in 35 of 1040 households (3.4%) in the Pacific Northwest between 2022-2024, with an estimated secondary attack rate of 15%. Sequences from within households clustered closely with 0 to 2 pairwise nucleotide differences (median 1) between cases 6-14 days apart (median 7).

## Main text

Enterovirus D68 (EV-D68) typically causes respiratory illness with symptoms such as difficulty breathing, fever, wheezing, and respiratory distress [1]. Interest in EV-D68 has grown in recent years due to associations with flaccid myelitis in children [2] and recent outbreaks in the United States and Europe [3]. In United States community studies, children under 16 years of age account for >80% of infections [4]. In congregate settings like homeless shelters, EV-D68 transmission was shown to occur primarily in adult, rather than family shelters [5]. There are no specific treatments or vaccines for non-polio enteroviruses, and the epidemiology of EV-D68, including risk factors for household transmission, are not well understood [6]. Very few studies have incorporated genomic sequencing to confirm transmission patterns or quantify viral evolution between transmission events [7,8].

Our study aimed to characterize the epidemiology and household transmission of EV-D68 using specimens and data collected as part of a community-based prospective cohort (CASCADIA [9]). The CASCADIA study included active surveillance for respiratory viruses among enrolled households with children and adults in metropolitan Seattle, Washington and Portland, Oregon, USA between June 2022 and March 2024 (Appendix). Participants completed surveys and self-collected nasal swabs on a weekly basis regardless of symptoms. Symptomatic swabs were tested by an array assay for 26 targets, EV-D68-specific RT-qPCR, and viral genome sequencing (Appendix).

After excluding 179 single-person households (see Appendix), 1,040 multi-person households were included in this analysis (Figure S1). A distinct EV-D68 index case was detected in 35 households (3.4%, Table 1). Potential secondary transmission, defined as detection of EV-D68 in a household contact 1-14 days after a distinct index case, was detected among 20% of households (n=7/35). The household secondary attack rate (SAR) for EV-D68 is estimated to be 15.0% (95% CI: 7.4-30.2%); and the median detection interval between index and secondary cases was 7 days (range 6-14 days). Among households with secondary transmission, fewer had children under 5 years of age (28.6% vs. 39.3%, n = 7 and 28); median age of the index case was higher (11 y vs. 9y); and more index cases had 2 or more acute respiratory illness symptoms (ARI, 100% vs. 68%), compared to households with unlikely secondary transmission. Households with secondary transmission had lower median household income (42.9% with income lower than the median income threshold vs. 78.6%) and a greater proportion of secondary contacts with comorbidities (70% vs. 55%) with compared to households with unlikely secondary transmission.

**Table 1.** Demographic characteristics of households with and without a secondary EV-D68 infection within 14 days of a distinct index case during the incident household illness episode, Washington and Oregon, United States, 2022-2024 (n=35)

|  |  | Potential<br>secondary<br>transmission<sup>1</sup> | Unlikely<br>secondary<br>transmission<sup>2</sup> | Total |
| --- | --- | --- | --- | --- |
| *Characteristic of household* |  | (n=7) | (n=28) | (N=35) |
| **Household density<sup>3</sup>** | 2-4 individuals | 6 (85.7%) | 24 (85.7%) | 30 (85.7%) |
|  | 5+ individuals | 1 (14.3%) | 4 (14.3%) | 5 (14.3%) |
| **Housing type** | House, condo, or townhouse | 7 (100%) | 27 (96.4%) | 34 (97.1%) |
|  | Missing | 0 | 1 (3.6%) | 1 (2.9%) |
| **Household enrollment** | Number of participants,<br>median [min, max] | 3 [2, 4] | 3 [2, 4] | 3 [2, 4] |
|  | No child <5 | 5 (71.4%) | 17 (60.7%) | 22 (62.9%) |
|  | Child <5, but no childcare | 1 (14.3%) | 2 (7.1%) | 3 (8.6%) |
|  | Child <5 in childcare<sup>4</sup> | 1 (14.3%) | 9 (32.1%) | 10 (28.6%) |
| **Income \$100,000+** | | 3 (42.9%) | 22 (78.6%) | 25 (71.4%) |
| **Smoker in household** |  | 1 (14.3%) | 2 (7.1%) | 3 (8.6%) |
| **Study site<sup>5</sup>** | KPNW | 4 (57.1%) | 12 (42.9%) | 16 (44.4%) |
|  | UW | 3 (42.9%) | 16 (57.1%) | 19 (54.3%) |
| *Characteristic of index case* |  | (n=7) | (n=28) | (N=35) |
| **Age** | Median [min, max] years | 11 [3, 34] | 9 [1, 48] | 9 [1, 48] |
|  | 6 months - 1 year | 0 (0.0%) | 2 (7.1%) | 2 (5.7%) |
|  | 2 - 4 years | 1 (14.3%) | 8 (28.0%) | 9 (25.7%) |
|  | 5 - 12 years | 3 (42.9%) | 8 (28.6%) | 11 (31.4%) |
|  | 13 - 50 years | 3 (42.9%) | 10 (35.7%) | 13 (37.1%) |
| **Gender** | Female | 3 (42.9%) | 17 (60.7%) | 20 (57.1%) |
|  | Male | 4 (57.1%) | 11 (39.3%) | 15 (42.9%) |
| **Race** | Asian | 0 (0.0%) | 1 (3.6%) | 1 (2.9%) |
|  | White | 3 (42.9%) | 22 (78.6%) | 25 (71.4%) |
|  | Multiracial | 3 (42.9%) | 4 (14.3%) | 7 (20.0%) |

<sup>1</sup> Potential secondary transmission defined as occurring when EV-D68 is detected in a secondary household member 1-14 days after index case.
<sup>2</sup> Unlikely secondary transmission defined as occurring when there is only 1 EV-D68 case detected in the household OR when there are 2+ EV-D68 cases in a household but secondary case detected >14 days after index case.
<sup>3</sup> Household density represents the number of household members regardless of enrollment in the study (thus, this may be greater than the number of individuals enrolled in the study if not all household members are enrolled).
<sup>4</sup> Daycare or school attendance among at least one child <5 years in the household as reported at enrollment
<sup>5</sup> KPNW = Kaiser Permanente Northwest; UW = University of Washington

|  |  |  |  |  |
| --- | --- | --- | --- | --- |
|  | Other | 1 (14.3%) | 1 (3.6%) | 2 (5.8%) |
| **Child attending school or daycare** |  | 5 (71.4%) | 18 (64.3%) | 23 (65.7%) |
| **Hispanic** |  | 0 (0.0%) | 3 (10.7%) | 3 (8.6%) |
| **Any smoking** |  | 0 | 0 | 0 |
| **Any comorbidities<sup>6</sup>** |  | 1 (14.3%) | 8 (28.6%) | 9 (25.7%) |
| **Masking in public** | Any | 7 (100%) | 23 (82.1%) | 30 (85.7%) |
|  | Never | 0 | 5 (17.9%) | 5 (14.4%) |
| **Relative cycle threshold (C<sub>rt</sub>)** | Median [Min, Max] | 18.8 [13.6, 21.8] | 16.0 [9.6, 22.5] | 17.3 [9.6, 22.5] |
|  | ≤ median (19.9 C<sub>rt</sub>) | 5 (71.4%) | 22 (78.6%) | 27 (77.1%) |
| **Duration of viral detection ≥ 1 week** |  | 0 | 0 | 0 |
| **Viral co-detections<sup>7</sup>** | Any | 7 (100%) | 26 (92.9%) | 33 (94.3%) |
|  | SARS-CoV-2 | 0 | 1 (3.6%) | 1 (2.9%) |
|  | Adenovirus | 1 (14.3%) | 0 (0%) | 1 (2.9%) |
|  | RSV | 0 | 1 (3.6%) | 1 (2.9%) |
|  | HPIV | 1 (14.3%) | 0 | 1 (2.9%) |
| **ARI symptom(s)<sup>8</sup>** | 2+ ARI symptoms | 7 (100%) | 19 (67.9%) | 26 (74.3%) |
|  | Cough and/or rhinorrhea | 7 (100%) | 28 (100%) | 35 (100%) |
| **Any care seeking during illness<sup>9</sup>** |  | 0 | 3 (10.7%) | 3 (8.6%) |
| **Any behavior change to reduce transmission in the air<sup>10</sup>** |  | 3 (42.9%) | 11 (39.3%) | 14 (40.0%) |
| **Any behavior change to reduce transmission on surfaces<sup>11</sup>** |  | 3 (42.9%) | 10 (35.7%) | 13 (37.1%) |
| ***Characteristics of household contacts*** |  | **(n=14)** | **(n=20)** | **(n=34)** |
| **Age** | Median [min, max] years | 42 [5, 48] | 26 [3, 49] | 38 [3, 49] |
|  | 2 - 4 years | 0 | 1 (5.0%) | 1 (2.9%) |
|  | 5 - 12 years | 3 (21.4%) | 8 (40.0%) | 11 (32.4%) |
|  | 13 - 50 years | 11 (78.6%) | 11 (55.0%) | 22 (64.7%) |
| **Female sex at birth** |  | 9 (64.3%) | 13 (65.0%) | 22 (64.7%) |
| **Gender** | Female | 9 (64.3%) | 12 (60.0%) | 22 (64.7%) |
|  | Male | 5 (35.7%) | 7 (35.0%) | 12 (35.3%) |
|  | Other | 0 | 1 (5.0%) | 1 (2.9%) |

<sup>6</sup> Comorbidities include asthma, chronic obstructive pulmonary disease (COPD) (including chronic bronchitis and emphysema), sleep apnea, heart disease, congenital heart disease, heart failure, down syndrome, hypertension (high blood pressure), diabetes (high blood sugar), liver condition, weak or failing kidneys, cancer or malignancy, arthritis, stroke, deep vein thrombosis (DVT) or pulmonary embolism (PE), sickle cell disease or thalassemia, weakened immune system, depression, anxiety, thyroid issues, or other health diagnosis.
<sup>7</sup> Rhinovirus target was detected in 32 (91.4%) samples, but is not shown here due to cross-reactivity of this target to EV-D68 and several other enterovirus species (Vi99990016\_po, Thermo Fisher)
<sup>8</sup> ARI, acute respiratory illness symptom reported within ±7 days of the individual EV-D68 illness episode's first positive specimen collection, including fever, cough, sore throat, shortness of breath, myalgia, and/or rhinorrhea. All individuals included reported at least one symptom.
<sup>9</sup> Self-reported care seeking defined as seeking health care (from a healthcare provider) during illness
<sup>10</sup> Includes masking, sleeping separately, covering cough/sneeze

|  |  |  |  |  |
| --- | --- | --- | --- | --- |
| Race | Asian | 2 (14.3%) | 1 (5.0%) | 3 (8.8%) |
|  | White | 10 (71.4%) | 16 (80.0%) | 26 (76.5%) |
|  | Multiracial | 2 (14.3%) | 3 (15.0%) | 5 (14.7%) |
| Hispanic |  | 0 | 2 (10.0%) | 2 (5.9%) |
| Any smoking |  | 1 (7.1%) | 2 (10.0%) | 3 (8.8%) |
| Any comorbidities<sup>12</sup> |  | 10 (71.4%) | 11 (55.0%) | 21 (61.8%) |
| Masking in public | Any | 13 (92.9%) | 20 (100.0%) | 33 (97.1%) |
|  | Never | 1 (7.1%) | 0 (0%) | 1 (2.9%) |

Samples from adults with either a primary or secondary infection tended to have lower minimum OpenArray C_rt_ values (mean 16.3, SD 4.27, n = 18) compared to children 6 months to 4 years of age (mean 17.5, SD 3.73, n = 11) and 5–17-year-olds (mean 17.1, SD 3.74, n = 15). Individuals who reported two or more ARI symptoms had similar Crt values (16.8, SD 4.12, n = 35) compared to those reporting a single symptom (16.8, SD 3.11, n = 9) (Figure S2).

High-quality EV-D68 whole genome sequences were recovered for two or more individuals within a household in a total of 11 households. All sequenced samples in this study (Table S1) fell within the B3 clade with other Genbank sequences from the United States and Europe; sequences from the same household clustered closely (Figure 1 and Figure S3). Among the 11 households, 6 had samples collected on the same day from different individuals (co-primary cases) and the within-household pairwise nucleotide (nt) distance between sequences ranged from 0 to 1 (median 0.5 nt). In 4 households, samples were collected 6-14 days apart and the within-household pairwise distance was 0-3 nt (median 1 nt). Sequences from one household with samples collected 148 days apart and sequences confirmed distinct introductions into the household.

**Figure 1.**
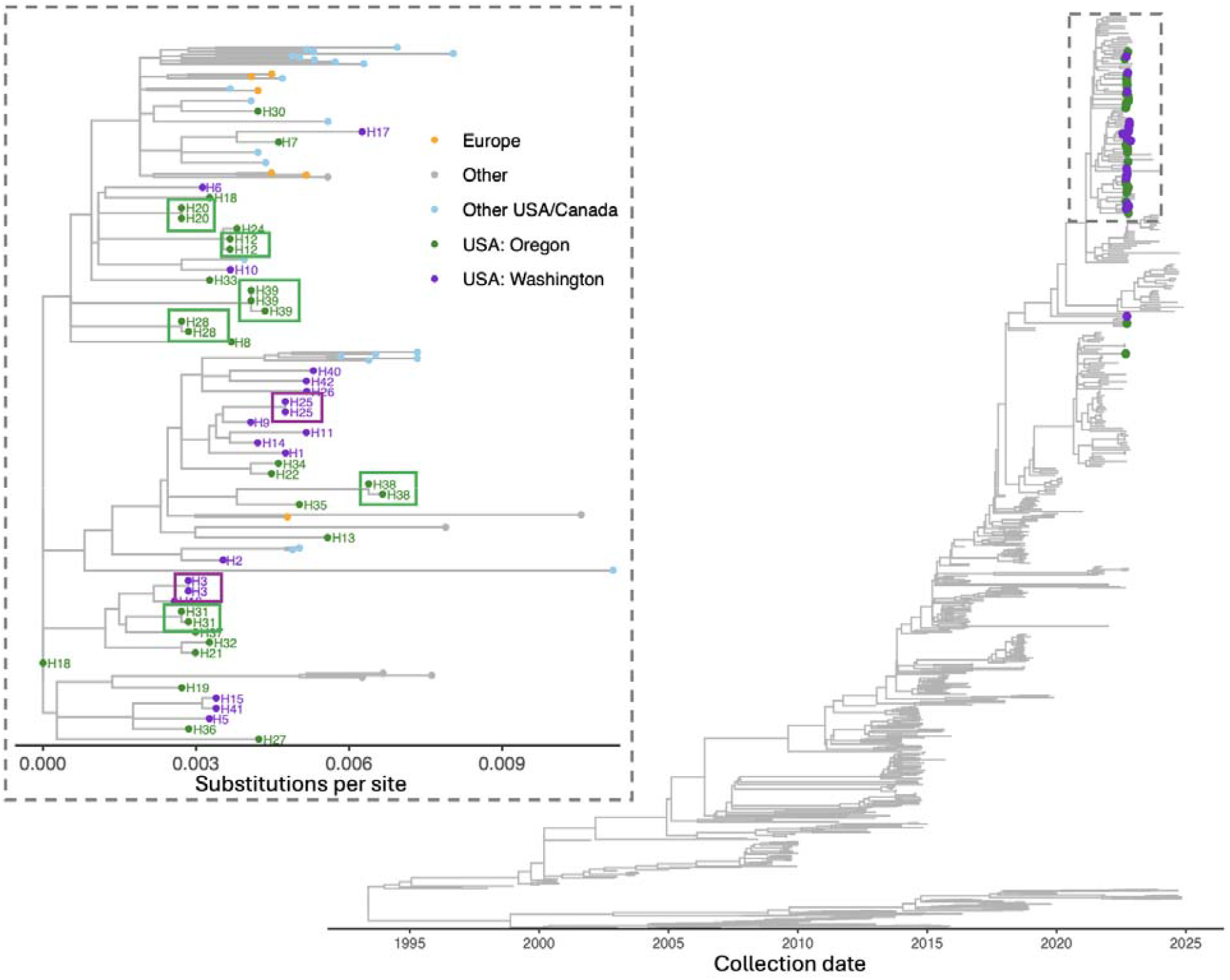
Maximum likelihood phylogenetic tree showing study samples in purple (Washington) or green (Oregon) and publicly available global sequences (no tip symbols). Inset figure shows sequences from the dashed region of the tree. Boxes around tips highlight close clustering of sequences from the same household.

As sequencing was performed using a hybridization capture-based method enriching for respiratory viruses (Appendix), a coinfecting pathogen (adenovirus, human parainfluenza virus 2, SARS-CoV-2) was identified in 3 samples along with EV-D68. The SARS-CoV-2 coinfection was in an adult, while the other two were in children under 5 years of age. We also sequenced two samples of Enterovirus C105 from a single household, which were identified as EV-D68 positive on the OpenArray due to cross-reactivity to non-EV-D68 enteroviruses described previously [5].

Overall, in this two-year community respiratory virus surveillance study, we found relatively few cases of EV-D68 in households compared to other respiratory viruses such as respiratory syncytial virus (RSV) [10]. The direction of transmission appears to be from school-aged children to adults, suggesting introductions to the household from children attending childcare or school. This work adds to the relatively scarce body of literature on enterovirus household transmission and confirms that in households with young children, EV-D68 is an uncommon source of respiratory viral illness as compared to other pathogens such as RSV, influenza virus, and SARS-CoV-2.

## Data Availability

All data produced in the present work are contained in the manuscript.

## Acknowledgements

The authors wish to acknowledge the following individuals for supporting this study. *Kaiser Permanente Center for Health Research*: Deralyn Almaguer, David Amy, Britt Ash, Allison Bianchi, Cassandra Boisvert, Stacy Bunnell, Joseph Cerizo, Evelin Coto, Phil Crawford, Robin Daily, Lantoria Davis, Stephen Fortmann, Kendall Frimodig, Lisa Fox, Holly Groom, Tarika Holness, Matt Hornbrook, Terry Kimes, Keelee Kloer, Dorothy Kurdyla, Bryony Melcher, John Ogden, Jennifer Rivelli, Katrina Schell, Emily Schield, Meagan Shaw, Martin Simer, Britta Torgrimson-Ojerio, Alexandra Varga, Mica Werner, Neil Yetz, Rebecca Ziebell; *University of Washington*: Ariana Magedson, Denise McCulloch, Natalie Lo, Kyle Luiten, Devon McDonald, Sarah Cox, Jenni Logue, Jean Mernaugh, Melissa MacMillan, Kat Hoffman, Grace Marshall, Jean Mernaugh, Daniel Nguyen, Zarna Marfatia, Amanda Casto, Chidozie Iwu, Julia Bennett, Jordan Opsahl, Kathryn McCaffrey, David Reinhart, Ben Cappodano, Sarah Heidl, Zack Acker, Lani Regelbrugge, Leslie Rodriguez-Salas, Ailyn Perez, Sean Ellis, Hanna Edgar; *Seattle Children’s Hospital:* Dallas Haws, Hanna Grioni, Josh Sanders, Irem Onalan, Laura Ostrina Restrepo.

## Author biography

Dr. Roychoudhury is Research Assistant Professor in the Department of Laboratory Medicine and Pathology at the University of Washington and Affiliate Investigator in the Vaccine and Infectious Disease Division at the Fred Hutchinson Cancer Center. Her primary research interests are pathogen genomics and mathematical models of viral evolution and host-pathogen interactions.

