## Appendix for "Household Transmission of Enterovirus D68 in Washington and Oregon, USA, 2022-2024"

**Supplemental Methods**

*Study Design and Population*

Study data were from the CASCADIA study, a community-based prospective cohort study among households of infants, children and adults in metropolitan Seattle, Washington and Portland, Oregon, USA [1]. From 1 June 2022 to March 2024, the study collected data using remote active surveillance of respiratory viruses among enrolled households, with at least weekly at-home collection of nasal swab specimens from participants with and without symptoms. Enrollment was open to people aged ≥6 months to 49 years living in the University of Washington (UW) catchment area (King, Pierce, and Snohomish counties) or the Kaiser Permanente catchment area (northern Oregon and southern Washington). Not all household members were required to enroll in the study. The study protocol was reviewed and approved by the Kaiser Permanente Inter-regional Institutional Review Board, with reliance from University of Washington and Seattle Children’s Research Institute (45 C.F.R. part 46.114; 21 C.F.R. part 56.114).

*Data collection*

Individuals were eligible for the CASCADIA study if they were between 6 months to 49 years of age during the time of enrollment. After consenting, participants were asked to complete an enrollment survey and an enrollment blood draw. Participants then completed weekly surveys and collected nasal swabs regardless of symptoms, and all swabs were tested for RSV, Influenza, and SARS-CoV-2. Nasal swabs from individuals who reported any new symptom (fever, chills, cough, shortness of breath, fatigue, sore throat, congestion or runny nose, nausea, vomiting, diarrhea, muscle or body aches, headache, and change in smell or taste, persistent pain/pressure in chest, pale/gray/blue lips skin or nail beds, and decreased activity and irritability/crankiness for young non-verbal children) within 72 hours from their weekly swab; as well as swabs that were inconclusive or positive for SARS-CoV-2, Influenza, or RSV also underwent testing on a multiplex PCR panel for 26 pathogens including EV-D68 as described below [1,2].

*PCR testing and viral sequencing*

Multiplex PCR testing was performed on selected nasal swab specimens using the OpenArray platform (ThermoFisher) with a custom panel containing 26 targets including EV-D68 as described previously [1,2]. Samples that were positive for the EV-D68 target were selected for this study. RNA was extracted using the MagNAPure 96 DNA and viral nucleic acid small volume kit (Roche Diagnostics), with 200μL input and 50μL elution. Where C_rt_ values are reported, the mean value for each sample was used from two replicates performed on the OpenArray assay.

Due to known cross-reactivity of the OpenArray EV-D68 target to other species including Coxsackievirus and Rhinovirus species [2], we performed PCR on a subset of samples to confirm EV-D68 positivity using the CDC Pan-Enterovirus D68 Real-Time RT-PCR Assay (primer and probe sequences in Table S2). PCR testing was run on the Applied Biosystem 7500 Fast system. PCR master mix was made using the Quanta qScript XLT One-Step RT-qPCR ToughMix (Quantabio).

For viral sequencing, extracted RNA was converted to double-stranded cDNA, purified by bead cleanup, enzymatically fragmented, end-repaired, indexed, amplified, and purified again using the QIAseq FX DNA Library Kit (Qiagen). Hybridization capture was performed using the QIAseq xHYB Viral Respiratory Panel (Qiagen) after pooling libraries by sample Crt values, with up to 6 samples in each pool. After overnight hybridization with biotinylated probes and subsequent washing to remove unbound fragments, enriched libraries were amplified and then purified by bead clean-up. Library fragment sizes were estimated by TapeStation 4200 D1000 (Agilent) and concentrations were measured by Qubit 4 Fluorometer (Invitrogen). Libraries passing quality control were sequenced on Illumina Novaseq 6000 or Nextseq 2000 instruments using a 2x150 read format. Consensus genomes were generated by using a custom bioinformatic pipeline (https://github.com/greninger-lab/revica) described previously [2]. This pipeline performs trimming of raw reads for quality, reference selection, and iterative mapping to generate a consensus genome. Sequencing data has been uploaded to NCBI BioProject PRJNA1029161 (Accessions available in Supplementary Table 1).

*Statistical analysis*

We summarized demographic, clinical, and behavioral characteristics of households, index cases, and household contacts. An **individual illness episode** was defined as the period within which an individual’s specimen(s) were PCR-positive for EV-D68 with ≤14 days separating any 2 positive specimens. A **household illness episode** was defined as a period within which ≥1 individual illness episode(s) occurred in member(s) of the same household with ≤14 days separating EV-D68 positive specimens in the household. As household transmission can only be assessed when there are multiple household members present, single-person households were excluded from the analysis. Within households with any EV-D68 infection, the **index case** was defined as the first household member with EV-D68 detected. **Co-primary index cases** were defined as two or more household members with the same date of EV-D68 detection. A **household contact** was defined as participants in the same household as an index case with a specimen(s) collected 1–14 days after the index case. **Potential secondary transmission** was defined as additional household contact(s) with EV-D68 detected 1-14 days after the date of detection of the index case(s). The **secondary attack rate** (SAR) was defined as the probability that an infection occurs among susceptible people within the same household [3]. We defined a **repeat detection** within the same individual as two detections of EV-D68 separated by >14 days with no positive results in the intervening period. **Detection interval** was defined as the number of days between the first positive specimen collection in the index case and the secondary case’s first EV-D68 positive specimen collection.

The primary outcome was secondary EV-D68 infection among household contacts within ≤14 days of an index case. The model predicting the secondary attack and 95% confidence intervals used an intercept-only generalized estimating equation (GEE) to account for household clustering, and included household contacts of a distinct index case. All analyses were conducted in R version 4.3.2

The household income cutoff of $100,000 was determined by median household income in the Seattle and Portland metropolitan areas. The median household income in 2022 was $134,600 for Seattle and $106,000 for Portland. Based on city-specific statistics and the income survey question structure, the decision was made to make a binary cutoff of $100,000 for models included in this analysis.

*Sequence analysis*

Trees were constructed using the Nextstrain platform [4]. Consensus sequences were aligned using augur after filtering and subsampling to include all study sequences and a random subset (5 per country-year-month) of contextual sequences downloaded from Genbank. Visualization was performed using auspice, and exported for annotation in ggtree [5]. Pairwise comparison of sequences within the household was performed by aligning sequences using MAFFT [6], masking sites with ambiguities, and counting the number of pairwise nucleotide differences across the genome using the ape package [7] in R version 4.4.2.

**Supplementary Figures**

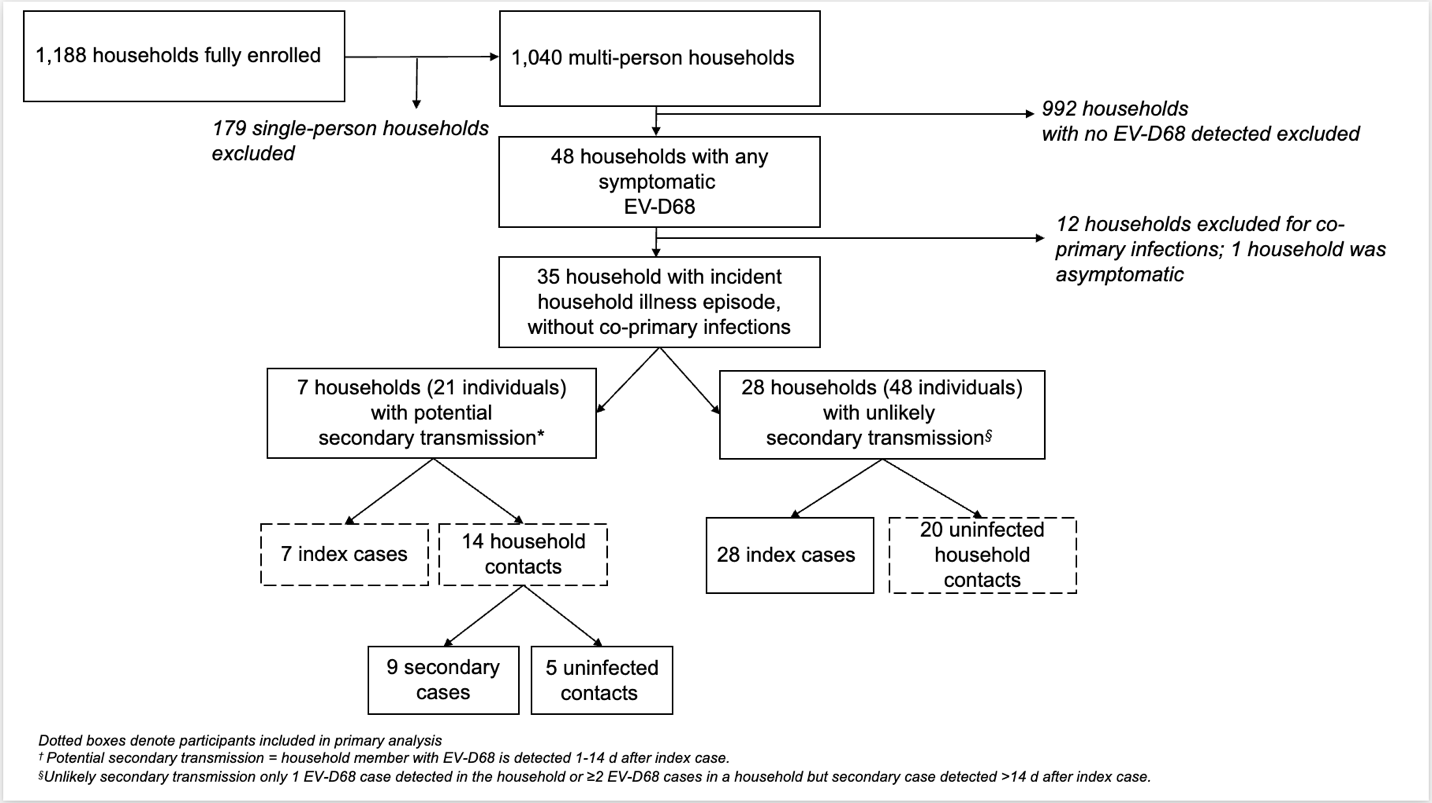

Figure S1. Study Flow Diagram

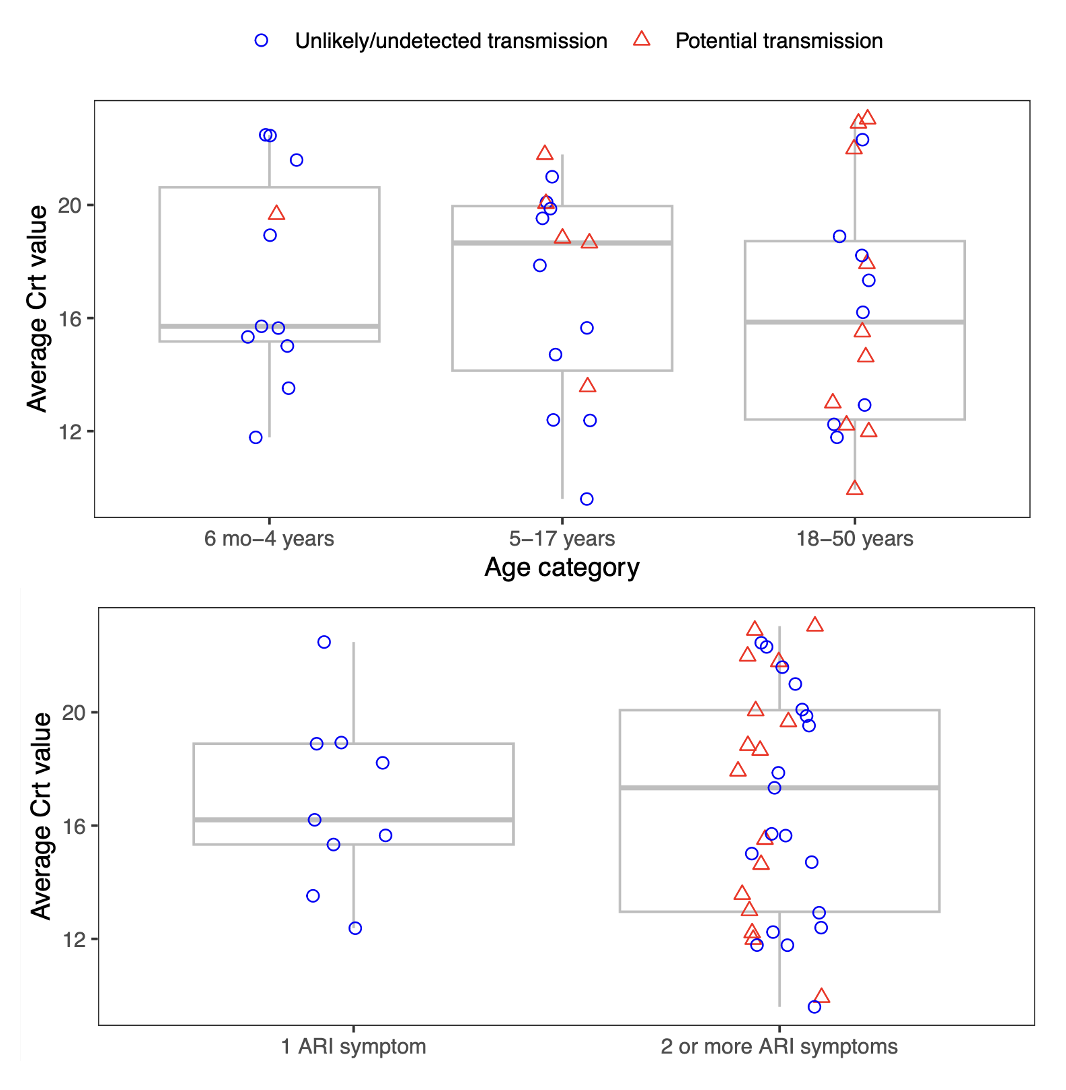

Figure S2. Comparing C_rt_ values across age groups (top panel) and by number of reported ARI symptoms (bottom panel).

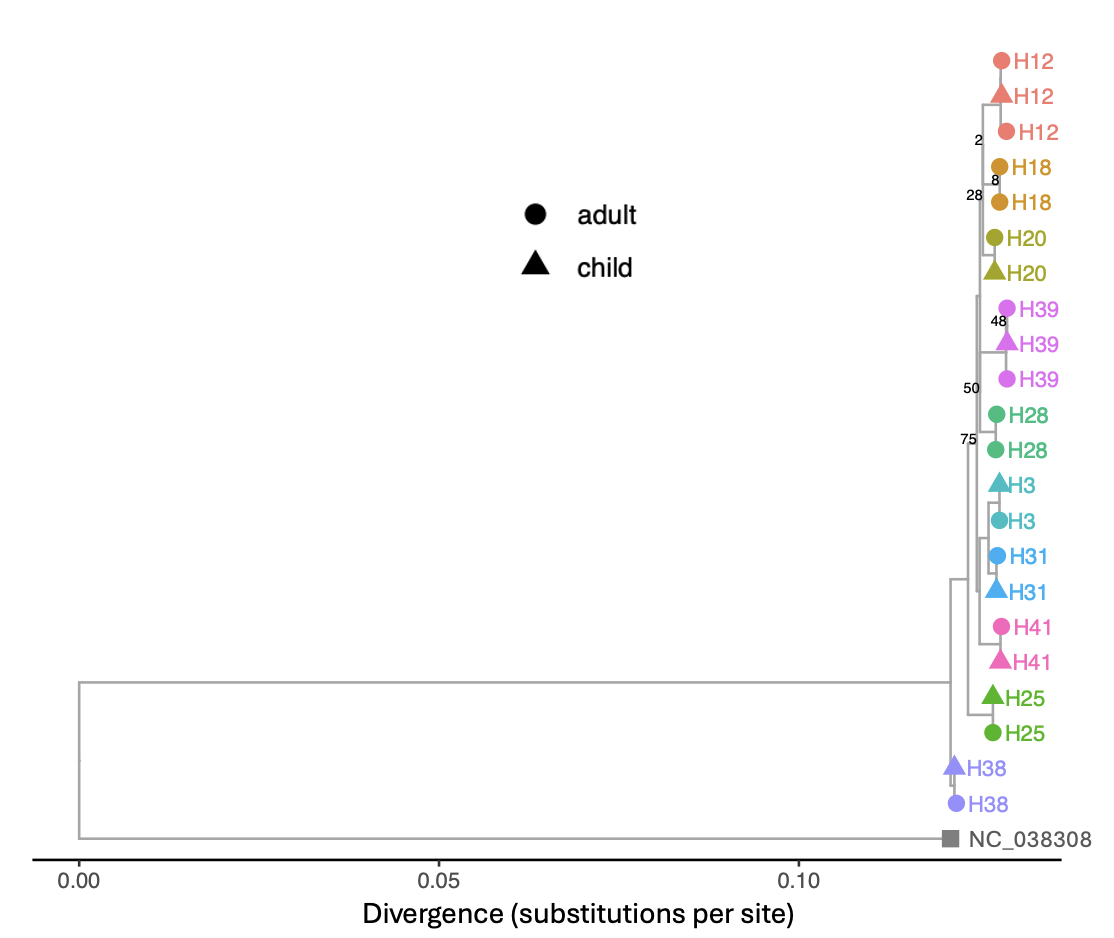

Figure S3. Maximum likelihood tree for sequences from 10 households with samples collected 0-14 days apart. Tip color indicates household, tip shape indicates whether the samples was from an adult or child. Bootstrap values shown only for nodes with <80% support.

**Supplementary Tables**

Table S1. Sequenced samples

| household | state | accession |
| --- | --- | --- |
| H1 | WA | PV448643 |
| H2 | WA | PV448634 |
| H3 | WA | PV448642 |
| H3 | WA | PV448638 |
| H4 | WA | PV448650 |
| H5 | WA | PV448647 |
| H6 | WA | PV448639 |
| H7 | OR | PV648971 |
| H8 | OR | PV648965 |
| H9 | WA | PV448644 |
| H10 | WA | PV448640 |
| H11 | WA | PV521983 |
| H12 | OR | PV660520 |
| H12 | OR | PV648964 |
| H13 | OR | PV660514 |
| H14 | WA | PV448646 |
| H15 | WA | PV448637 |
| H16 | WA | PV448648 |
| H17 | WA | PX048941 |
| H18 | OR | PV660513 |
| H18 | OR | PV648972 |
| H19 | OR | PV648966 |
| H20 | OR | PV660519 |
| H20 | OR | PV648957 |
| H21 | OR | PV660517 |
| H22 | OR | PV660515 |
| H23 | OR | PV648961 |
| H24 | OR | PV660516 |
| H25 | WA | PX048942 |
| H25 | WA | PV448636 |
| H26 | WA | PV448645 |
| H27 | OR | PV660521 |
| H28 | OR | PV648967 |
| H28 | OR | PV648963 |
| H29 | OR | PV648958 |
| H30 | OR | PV648956 |
| H31 | OR | PV660511 |
| H31 | OR | PV648969 |
| H32 | OR | PV660523 |
| H33 | OR | PV660512 |
| H34 | OR | PV660522 |
| H35 | OR | PV648973 |
| H36 | OR | PV648962 |
| H37 | OR | PV648959 |
| H38 | OR | PV648970 |
| H38 | OR | PV648960 |
| H39 | OR | PX048940 |
| H39 | OR | PV660518 |
| H39 | OR | PV648968 |
| H40 | WA | PV448635 |
| H41 | WA | PV448641 |
| H42 | WA | PV448649 |
| H43 | WA | pending |
| H44 | WA | pending |
| H19 | OR | pending |
| H19 | OR | pending |
| H47 | OR | pending |
| H39 | OR | pending |
| H41 | WA | pending |
| H41 | WA | pending |
| H42 | WA | pending |
| H45 | OR | pending |
| H46 | OR | pending |

Table S2. EV-D68 PCR primers and probe sequences

| Primer/Probe | Sequence |
| --- | --- |
| AN993 | 5’ GGAATAAATCCAGCNGAYACNAT 3’ |
| AN995 | 5’ CCACGCTTTTATRTGYTTNGGYTTCAT 3’ |
| AN992 | 5’ FAM GARCAYCARCCARTTGGTTTCACAGTGAC BHQ1 3’ |
